# Analysis of humoral immunity against emerging SARS-CoV-2 variants: a population-based prevalence study in Yokohama, Japan

**DOI:** 10.1101/2022.03.26.22272766

**Authors:** Atsushi Goto, Kei Miyakawa, Izumi Nakayama, Susumu Yagome, Juan Xu, Makoto Kaneko, Norihisa Ohtake, Hideaki Kato, Akihide Ryo

## Abstract

**Background:** Little is known about the population prevalence of antibodies against emerging immune escape variants of SARS-CoV-2.

**Methods:** A population-based prevalence study was conducted in Yokohama City, the most populous municipality of Japan. Quantitative measurements of immunoglobulin G against SARS-CoV-2 spike protein (SP-IgG) and qualitative measurements of neutralization antibodies against the Omicron BA.1 and BA.2 variants were performed.

**Results:** Of 6,000 randomly selected residents aged 20–74, 1,277 participated in the study during a period from January 30 to February 28, 2022. Of them, 3% had prior diagnosis of COVID-19, 96% received at least two-doses of SARS-CoV-2 vaccines, and 94% were positive for SP-IgG. The positive rates of neutralizing antibodies were 28% to Omicron BA.1 and BA.2 variants in a random sample of 10% of participants (n=123) and 100% to BA.1 and BA.2 among participants who received the third vaccination at least 7 days before (n=66).

**Conclusions:** In this population-based prevalence study in Japan, most had SP-IgG antibodies but the overall neutralizing antibody positive rate was 28% against the Omicron BA.1 and BA.2 variants. The population-level insufficient humoral immunity against the Omicron variants may explain the outbreak of COVID-19 during this period in Japan.

## INTRODUCTION

The coronavirus disease 2019 (COVID-19), first reported in Wuhan, Hubei Province, China, in December 2019 rapidly spread globally after March 2020 to become pandemic. As of March 2022, the most effective strategy against COVID-19 is vaccination of the population. Severe acute respiratory coronavirus-2 (SARS-CoV-2) vaccines, such as Pfizer-BioNTech BNT162b2, Moderna mRNA-1273, and AstraZeneca ChAdOx1-S, are now being used worldwide. Internationally, COVID-19 Vaccines Global Access (COVAX) [1] is leading a global collaboration to accelerate the development and manufacture of COVID-19 vaccines and guarantee fair and equitable access to every country in the world.

Vaccination against COVID-19 was almost completed by the end of 2021 for those who wished to receive two doses of the vaccine in Japan. However, the antibody titers decrease 6 months to 1 year after vaccination, and even individuals who have received two doses of the vaccine can be infected (i.e., breakthrough infection) [2, 3]. Under these circumstances, the third vaccination (booster shot) was administered in December 2021 after an interval of 6–8 months from the completion of the second vaccination. In Japan, approximately 80% of the total population have received at least two doses of the vaccine against SARS-CoV-2, and approximately 33% have received the third shot of the vaccination as of March 18, 2022 [4], which is similar to vaccination rates in other high-income countries [5]. However, there is an emerging concern that the variants of concern (VOC), particularly the Omicron variant (B.1.1.529 or BA.1), would escape the antibodies elicited by vaccination against SARS-CoV-2 [6, 7]. In fact, Japan has been experiencing a resurgence of COVID-19 cases [8] with the Omicron variant becoming dominant since January 2022[9], despite most of the adult population have completed two doses of vaccines [10].

From public health perspectives, evaluation of a population-level immunity to SARS-CoV-2, including the Omicron BA.1 and the newly emerged Omicron sub-lineage BA.2 (B.1.1.529.2) variants, is essential to facilitate evidence-informed decision-making regarding COVID-19. However, only a few studies have reported the population-level prevalence of antibodies against SARS-CoV-2 [11] and VOC, such as Delta (B.1.617.2) and Omicron BA.1 [12]. Moreover, to the best of our knowledge, there is a lack of data on population immunity against the newly emerged variant, the sub-lineage BA.2 of the Omicron variant, at present. Since Omicron BA.2 has begun to dominate globally[13], an investigation of the immunity against BA.2 at a population level is urgently warranted.

Therefore, in this population-based prevalence study in Yokohama City, the most populous municipality of Japan, we evaluated population-level humoral immunity against emerging immune escape variants such as Omicron BA.1 and BA.2. The survey was undertaken from January 30 to February 28, 2022, during the middle of the sixth wave of COVID-19 in Japan, which was driven by the emergence of the Omicron BA.1.

## METHODS

### Study population

In total, 6,000 residents were randomly selected from a Japanese population aged 20–74 year, from Yokohama City, Japan. With a population of approximately 3.7 million, Yokohama City is the most populated basic municipality in Japan and is located in Kanagawa Prefecture next to Tokyo. The study invitations were mailed to these residents in January 2022. Among those, 1,277 individuals (546 men and 731 women; response rate: 21.3%), who did not have a confirmed diagnosis of COVID-19 within 2 weeks of the study entry, participated in this study during a period from January 30 to February 28, 2022. The participation rate was relatively high among middle-aged (45-59) women with response rate being 30% or more (Supplemental Figure 1). All participants provided written informed consent. The study was approved by the Institutional Review Board of the Yokohama City University.

### Measurements

Each participant provided approximately 7 ml of blood samples and completed a questionnaire with questions regarding their prior COVID-19 diagnosis, vaccination against SARS-CoV-2, and lifestyle and social factor at study entry. Participants who provided incomplete answers, such as incorrect vaccination date, were supplemented with an additional questionnaire or underwent telephone interview. Therefore, there were not missing data on variables listed in Table 1.

**Table 1.**
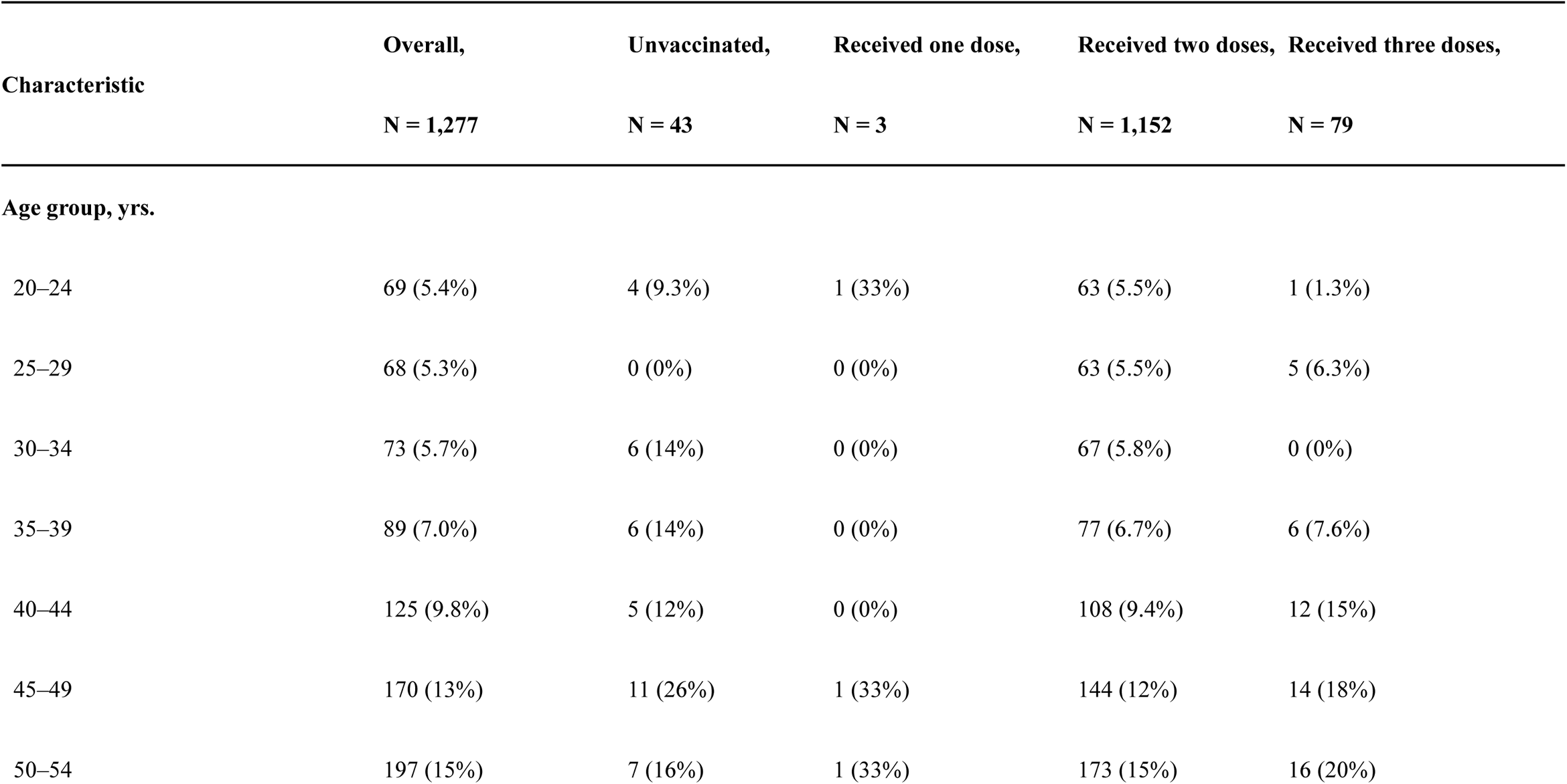
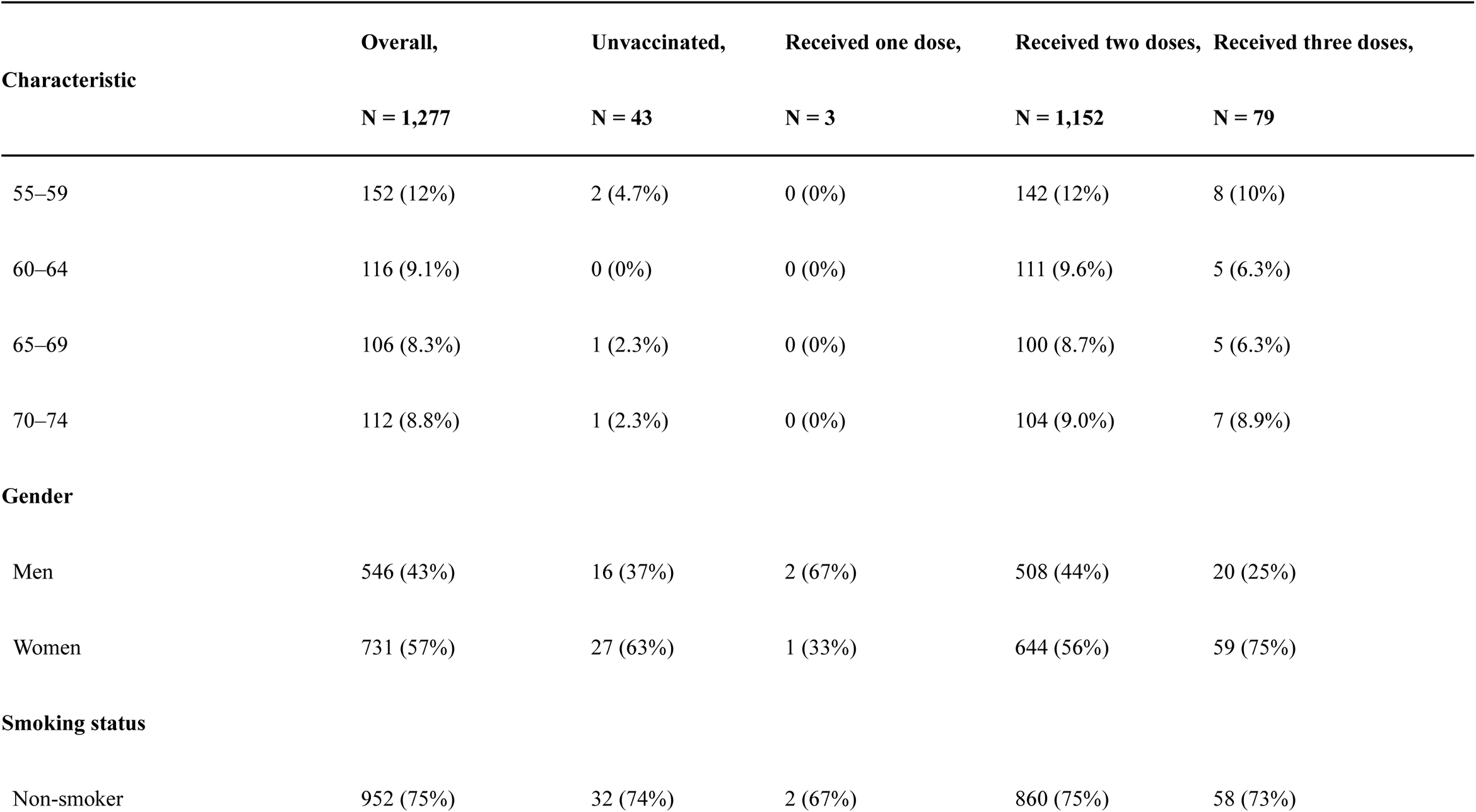
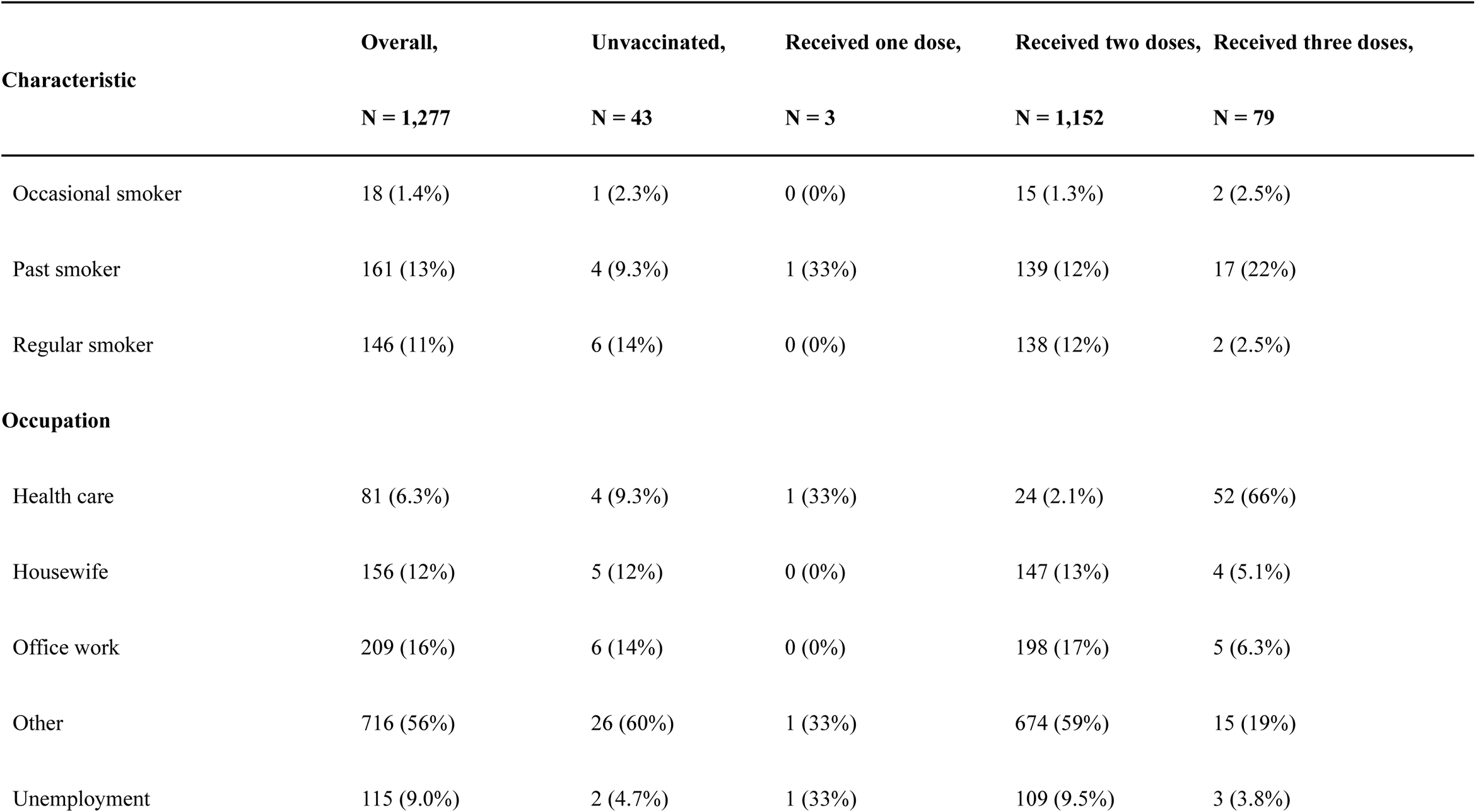
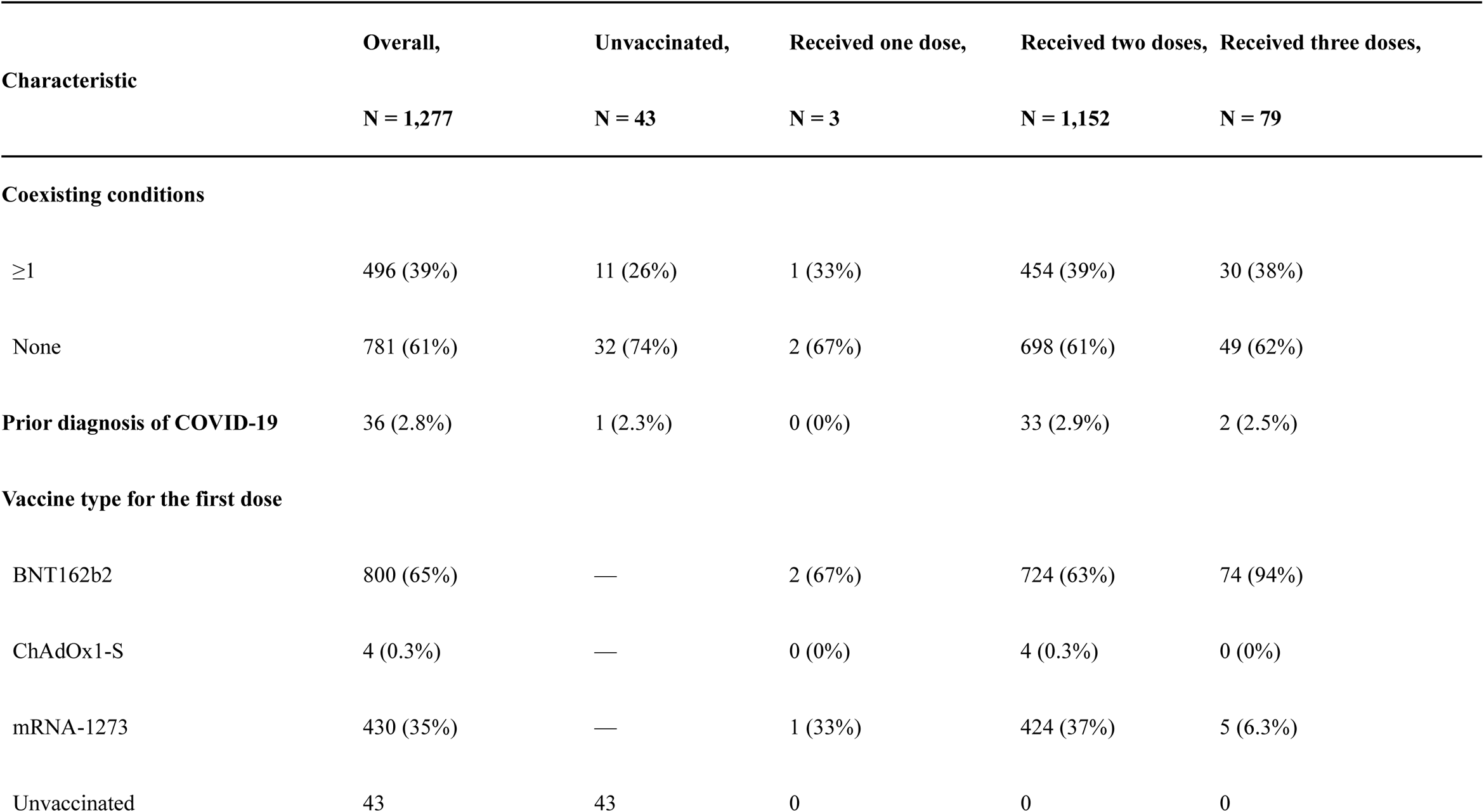
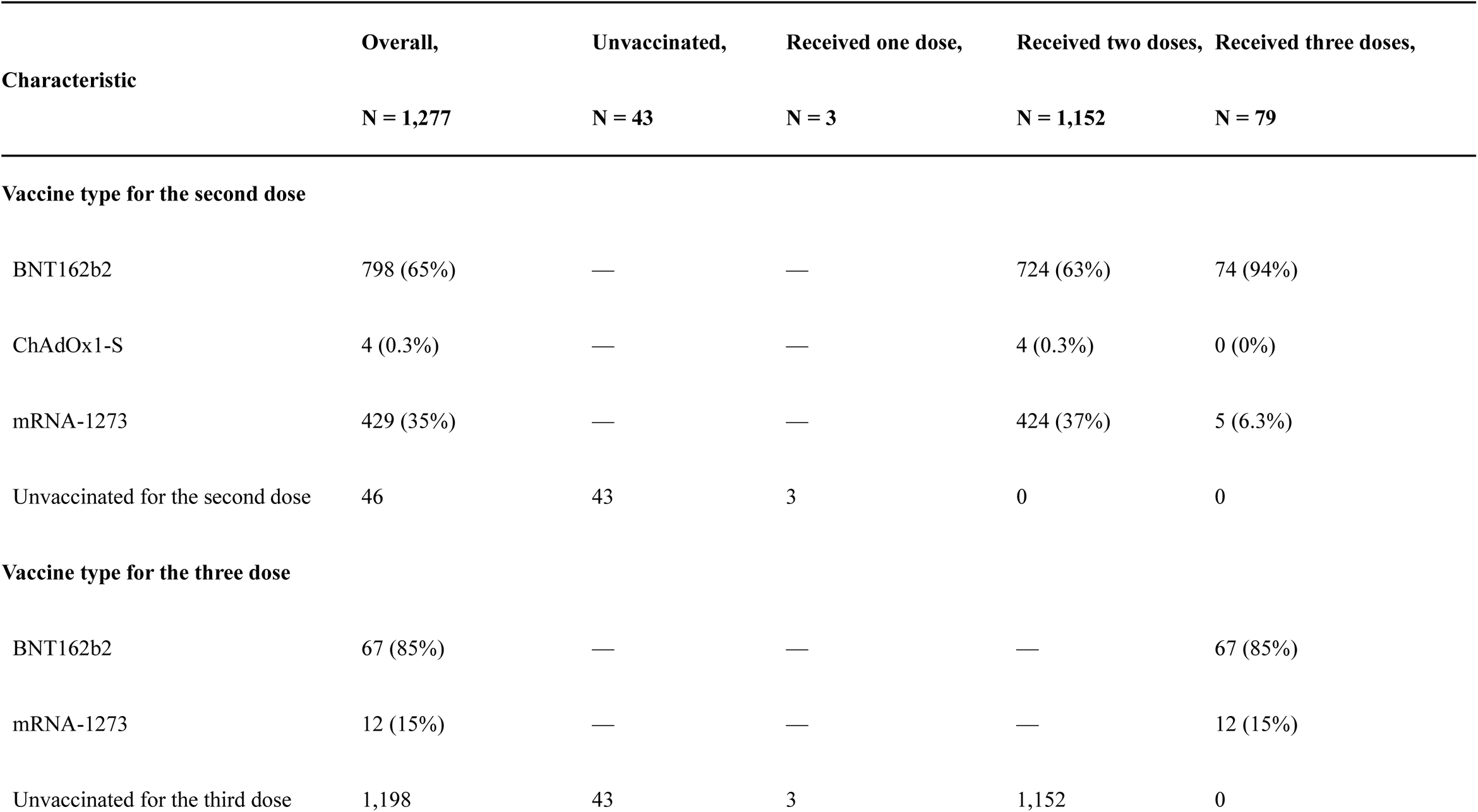
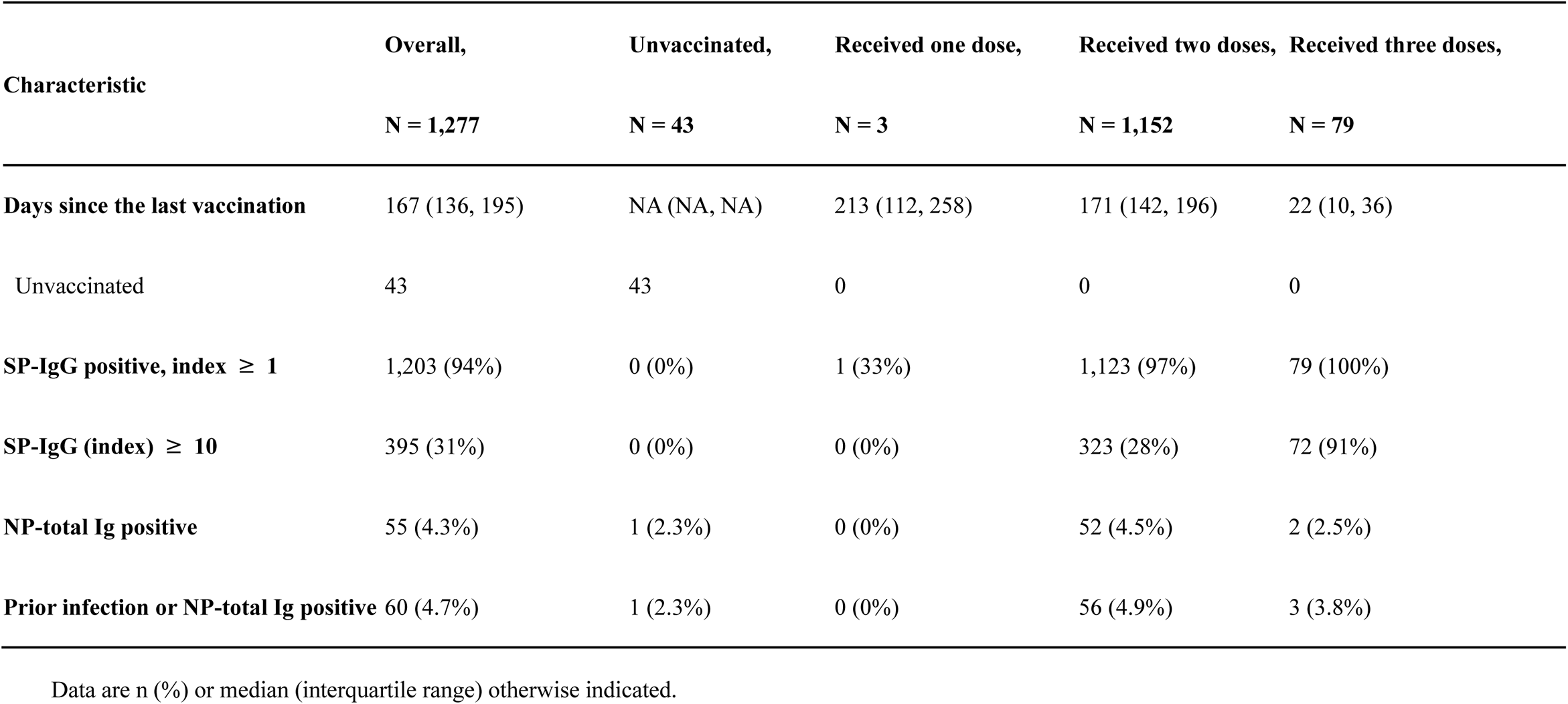
Baseline characteristics according to vaccination status (N = 1,277)

| **Characteristic** | **Overall,**<br>**N = 1,277** | **Unvaccinated,**<br>**N = 43** | **Received one dose,**<br>**N = 3** | **Received two doses,**<br>**N = 1,152** | **Received three doses,**<br>**N = 79** |
| --- | --- | --- | --- | --- | --- |
| **Age group, yrs.** |  |  |  |  |  |
| 20–24 | 69 (5.4%) | 4 (9.3%) | 1 (33%) | 63 (5.5%) | 1 (1.3%) |
| 25–29 | 68 (5.3%) | 0 (0%) | 0 (0%) | 63 (5.5%) | 5 (6.3%) |
| 30–34 | 73 (5.7%) | 6 (14%) | 0 (0%) | 67 (5.8%) | 0 (0%) |
| 35–39 | 89 (7.0%) | 6 (14%) | 0 (0%) | 77 (6.7%) | 6 (7.6%) |
| 40–44 | 125 (9.8%) | 5 (12%) | 0 (0%) | 108 (9.4%) | 12 (15%) |
| 45–49 | 170 (13%) | 11 (26%) | 1 (33%) | 144 (12%) | 14 (18%) |
| 50–54 | 197 (15%) | 7 (16%) | 1 (33%) | 173 (15%) | 16 (20%) |

| Characteristic | Overall, | Unvaccinated, | Received one dose, | Received two doses, | Received three doses, |
| --- | --- | --- | --- | --- | --- |
|  | N = 1,277 | N = 43 | N = 3 | N = 1,152 | N = 79 |
| 55–59 | 152 (12%) | 2 (4.7%) | 0 (0%) | 142 (12%) | 8 (10%) |
| 60–64 | 116 (9.1%) | 0 (0%) | 0 (0%) | 111 (9.6%) | 5 (6.3%) |
| 65–69 | 106 (8.3%) | 1 (2.3%) | 0 (0%) | 100 (8.7%) | 5 (6.3%) |
| 70–74 | 112 (8.8%) | 1 (2.3%) | 0 (0%) | 104 (9.0%) | 7 (8.9%) |
| **Gender** |  |  |  |  |  |
| Men | 546 (43%) | 16 (37%) | 2 (67%) | 508 (44%) | 20 (25%) |
| Women | 731 (57%) | 27 (63%) | 1 (33%) | 644 (56%) | 59 (75%) |
| **Smoking status** |  |  |  |  |  |
| Non-smoker | 952 (75%) | 32 (74%) | 2 (67%) | 860 (75%) | 58 (73%) |
| Occasional smoker | 18 (1.4%) | 1 (2.3%) | 0 (0%) | 15 (1.3%) | 2 (2.5%) |
| Past smoker | 161 (13%) | 4 (9.3%) | 1 (33%) | 139 (12%) | 17 (22%) |
| Regular smoker | 146 (11%) | 6 (14%) | 0 (0%) | 138 (12%) | 2 (2.5%) |
| **Occupation** |  |  |  |  |  |
| Health care | 81 (6.3%) | 4 (9.3%) | 1 (33%) | 24 (2.1%) | 52 (66%) |
| Housewife | 156 (12%) | 5 (12%) | 0 (0%) | 147 (13%) | 4 (5.1%) |
| Office work | 209 (16%) | 6 (14%) | 0 (0%) | 198 (17%) | 5 (6.3%) |
| Other | 716 (56%) | 26 (60%) | 1 (33%) | 674 (59%) | 15 (19%) |
| Unemployment | 115 (9.0%) | 2 (4.7%) | 1 (33%) | 109 (9.5%) | 3 (3.8%) |

| Characteristic | Overall,<br>N = 1,277 | Unvaccinated,<br>N = 43 | Received one dose,<br>N = 3 | Received two doses,<br>N = 1,152 | Received three doses,<br>N = 79 |
| --- | --- | --- | --- | --- | --- |
| **Coexisting conditions** |  |  |  |  |  |
| ≥1 | 496 (39%) | 11 (26%) | 1 (33%) | 454 (39%) | 30 (38%) |
| None | 781 (61%) | 32 (74%) | 2 (67%) | 698 (61%) | 49 (62%) |
| **Prior diagnosis of COVID-19** | 36 (2.8%) | 1 (2.3%) | 0 (0%) | 33 (2.9%) | 2 (2.5%) |
| **Vaccine type for the first dose** |  |  |  |  |  |
| BNT162b2 | 800 (65%) | — | 2 (67%) | 724 (63%) | 74 (94%) |
| ChAdOx1-S | 4 (0.3%) | — | 0 (0%) | 4 (0.3%) | 0 (0%) |
| mRNA-1273 | 430 (35%) | — | 1 (33%) | 424 (37%) | 5 (6.3%) |
| Unvaccinated | 43 | 43 | 0 | 0 | 0 |
| **Vaccine type for the second dose** |  |  |  |  |  |
| BNT162b2 | 798 (65%) | — | — | 724 (63%) | 74 (94%) |
| ChAdOx1-S | 4 (0.3%) | — | — | 4 (0.3%) | 0 (0%) |
| mRNA-1273 | 429 (35%) | — | — | 424 (37%) | 5 (6.3%) |
| Unvaccinated for the second dose | 46 | 43 | 3 | 0 | 0 |
| **Vaccine type for the three dose** |  |  |  |  |  |
| BNT162b2 | 67 (85%) | — | — | — | 67 (85%) |
| mRNA-1273 | 12 (15%) | — | — | — | 12 (15%) |
| Unvaccinated for the third dose | 1,198 | 43 | 3 | 1,152 | 0 |
| **Days since the last vaccination** | 167 (136, 195) | NA (NA, NA) | 213 (112, 258) | 171 (142, 196) | 22 (10, 36) |
| Unvaccinated | 43 | 43 | 0 | 0 | 0 |
| **SP-IgG positive, index $\geq 1$** | 1,203 (94%) | 0 (0%) | 1 (33%) | 1,123 (97%) | 79 (100%) |
| **SP-IgG (index) $\geq 10$** | 395 (31%) | 0 (0%) | 0 (0%) | 323 (28%) | 72 (91%) |
| **NP-total Ig positive** | 55 (4.3%) | 1 (2.3%) | 0 (0%) | 52 (4.5%) | 2 (2.5%) |
| **Prior infection or NP-total Ig positive** | 60 (4.7%) | 1 (2.3%) | 0 (0%) | 56 (4.9%) | 3 (3.8%) |

Data are n (%) or median (interquartile range) otherwise indicated.

Following prior publications [14–18], IgG antibodies against spike protein (SP-IgG) or total Ig antibodies against nucleocapsid protein (NP-total Ig) were measured using the commercial chemiluminescent enzyme immunoassay (AIA-CL SARS-CoV-2 SP-IgG and NP-total Ig antibody detection reagents, Tosoh, Japan) to assess prior infection (NP-total Ig) or vaccination against SARS-CoV-2 (SP-IgG or NP-total Ig). An index of ≥ 1.0 for these antibodies were considered positive according to the manufacturer’s instructions. The limit of detection (LOD) for SP-IgG and NP-total Ig was 0.1, and those with values below LOD were assigned a value of 0.05 for statistical analyses. For the assessment of neutralizing antibodies, a rapid qualitative neutralizing assay was performed as previously reported [15] using the spikes of D614G as a reference and three variants (Delta, Omicron BA.1, and BA.2) among a random sample of 10% (n=123) of the total study population and 79 participants who received the booster shot of the SARS-CoV-2 vaccine. Sera were assayed using the rapid qualitative neutralization test (hiVNT) with the HiBiT-tagged virus-like particle carrying SARS-CoV-2 spike. At a fixed serum sample dilution of 1 in 20, if luminescence signal inhibition exceeds 40%, a sample is considered to possess neutralizing activity equivalent to the neutralizing titer of 51 or more in the HIV-based pseudovirus method [15].

### Statistical Analysis

We computed an overall crude antibody prevalence rate and its 95% confidence interval (CI) for the positivity of SP-IgG antibody as defined as an index of ≥ 1.0 and a weighted prevalence rate and 95% CI to adjust for differences in participation rate across age and sex and represent the overall population aged 20–74 of Yokohama City. For the weighted prevalence rate, we calculated the participation rate by sex and age group in 5-year increments. Subsequently, we computed the weighted prevalence rate and its 95% CI with weights with the reciprocal of the participation rates, using the survey package [19] in R. For the crude prevalence, the Clopper–Pearson method was used to estimate CI using the binom.test function in R.

Characteristics such as age, number of vaccinations (none, one, two, or three does), and number of days since vaccination in relation to SP-IgG index were investigated by stratification and linear regression analyses, after excluding those with prior COVID-19 diagnosis or positive NP-total Ig, which indicates prior infection with SARS-CoV-2. In regression analyses, after excluding those with prior COVID-19 diagnosis or positive NP-total Ig, log-10 transformed SP-IgG index was regressed on age or days since the last vaccination among those who completed two doses of vaccines.

Among randomly selected 10% subpopulations (n=123) of the total participants, we examined the positive rates (95% CIs using the Clopper–Pearson method) for neutralization antibodies against the Delta, Omicron BA.1, and BA.2 variants with D614G as a reference strain. In addition, we examined the positive rates neutralization antibodies against these variants among those who received the three doses of vaccines against SARS-CoV-2 at least 7 days before, after excluding those with prior COVID-19 diagnosis or positive NP-total Ig.

All statistical analyses were performed with R version 4.1.2 (The R Foundation for Statistical Computing).

## RESULTS

### Baseline characteristics

Among 1,277 participants (546 men, 43%; 731 women, 57%), most had received two doses (but not the third dose) of vaccination, accounting for 90.2%; 6.2% had completed the third (booster) dose, 0.2% had received the first dose, and 3.4% had not been vaccinated (Table 1). Among them, 2.8% had been previously diagnosed with COVID-19 and 4.3% had tested positive for NP-total Ig, and 4.7% (= 60/1,277) were considered to have prior COVID-19.

### Prevalence of SP-IgG positive

Approximately 94% of the participants tested positive for SP-IgG antibodies (weighted prevalence rate: 94.2%; 95% CI: 93.0–95.4%; crude prevalence rate: 94.2%; 95% CI: 92.8–95.4%; not shown in tables).

### SP-IgG index values according to numbers of vaccination and types of vaccines

Among 1,217 individuals with no history of infection and who tested negative for NP-Ig total, the values of SP-IgG index were the highest among those who completed three doses of vaccine (geometric mean titer [GMT]: 32.2; 95% CI: 26.6–38.9), followed by two doses (GMT: 5.85; 95% CI: 5.55–6.16), one dose (GMT: 0.6; 95%CI: 0.2–1.8), and unvaccinated (GMT: 0.11; 95%CI: 0.01–0.13) (Figure 1A). Further, those who received two doses of Moderna mRNA-1273 (GMT: 9.1; 95% CI: 8.6– 9.6) tended to have a higher SP-IgG index than those who had received two doses of the Pfizer-BioNTech BNT162b2 vaccine (GMT 4.0; 95% CI: 3.8-4.3) (Figure 1B).

**Figure 1.**
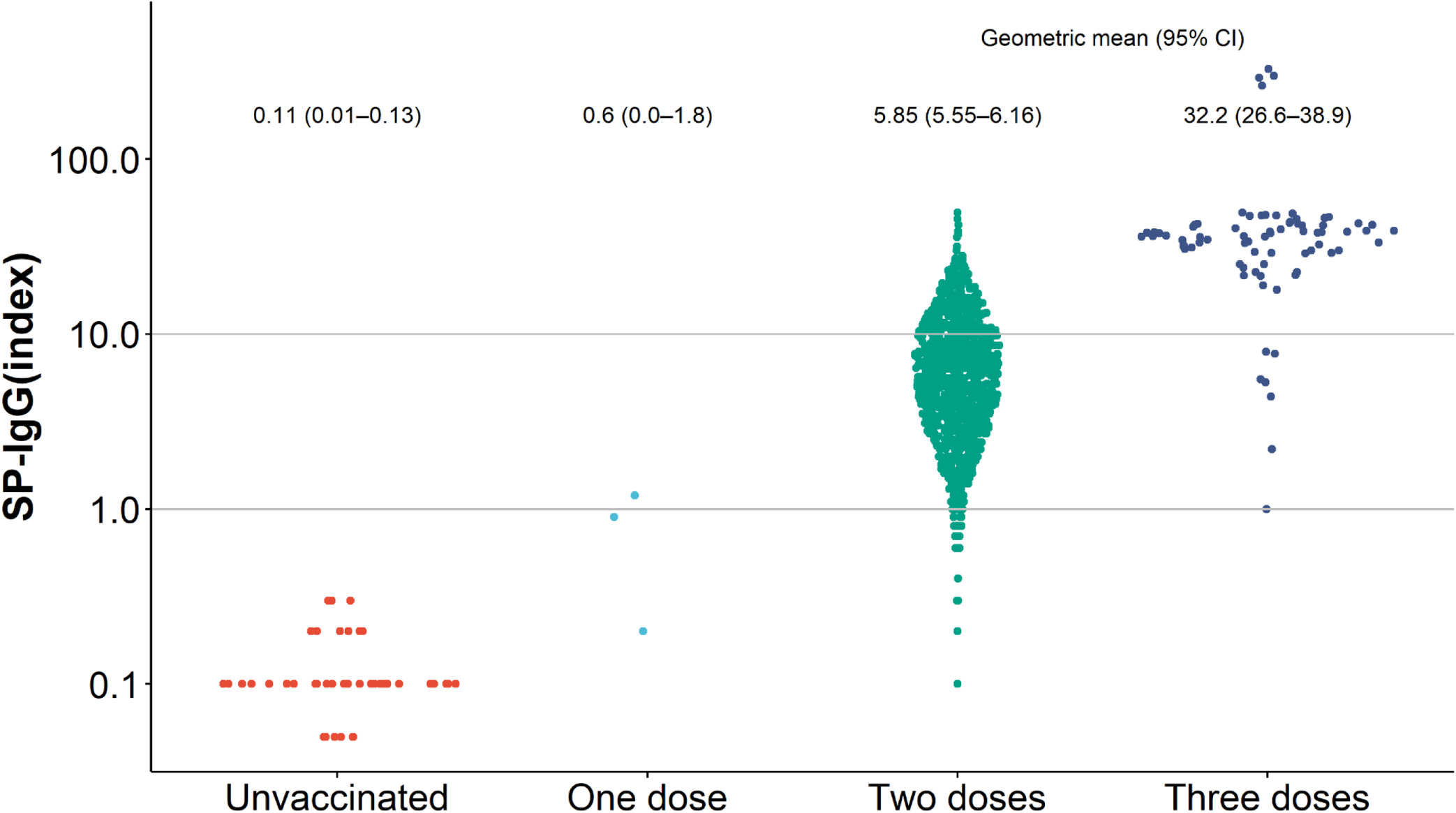
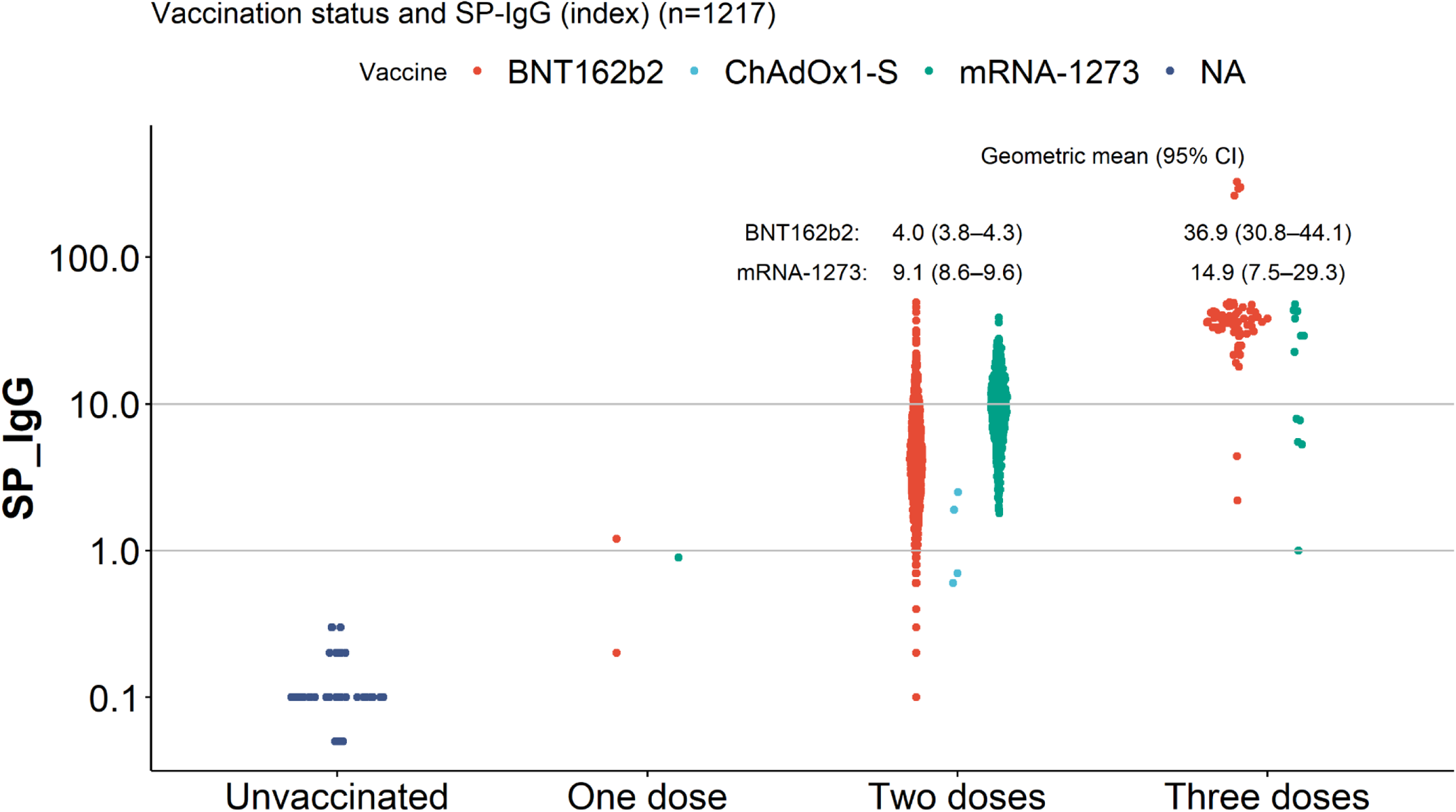
The SP-IgG index according to vaccination status and vaccine types Geometric mean of SP-IgG index (95% confidence interval) are shown. Panel A shows the distribution of SP-IgG index according to vaccination status (n=1,217). Panel B shows the distribution of SP-IgG index according to vaccination status and types of vaccines (n=1,217).

### SP-IgG index and characteristics among participants with two doses of vaccines

In 1,092 individuals with no history of infection and who tested negative for NP-Ig total and received two doses of Pfizer-BioNTech BNT162b2 or Moderna mRNA-1273 vaccine, as more days elapsed since the second dose, the lower the SP-IgG index tended to be (Figure 2A). At any number of days, the SP-IgG index values were higher among those who received the Moderna mRNA-1273 vaccine than among those who received the Pfizer-BioNTech BNT162b2 vaccine. In addition, the older the age, the more SP antibody titers tended to be low; this trend was prominent for the BNT162b2 vaccine (Figure 2B). In 1,092 individuals with no history of infection and who tested negative for NP-total Ig and received two doses of the BNT162b2 or mRNA-1273 vaccine, those who received the BNT162b2 vaccine tended to have more adverse reactions, such as fever, than those who received the mRNA-1273 vaccine (Supplemental Figure 2).

**Figure 2.**
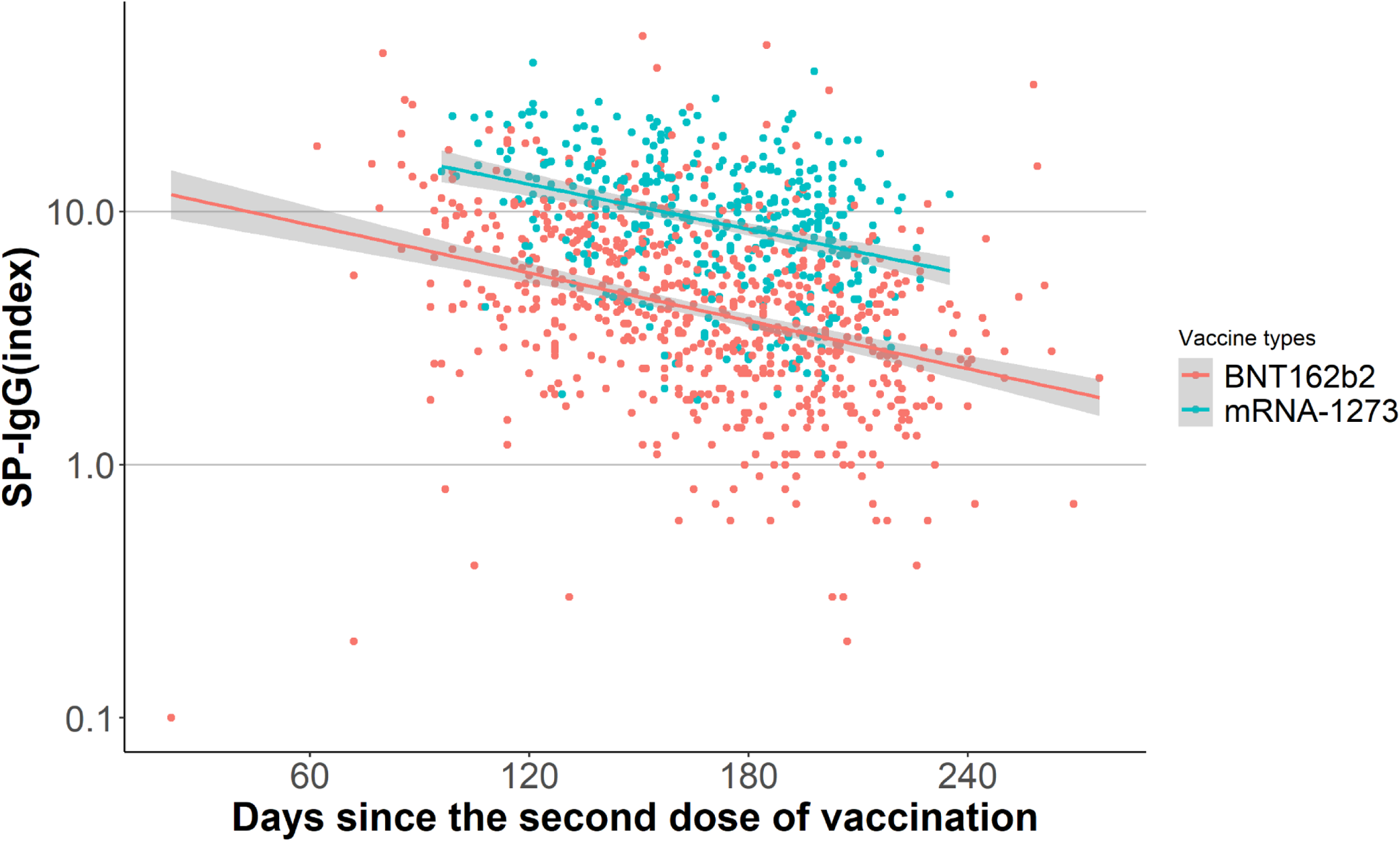
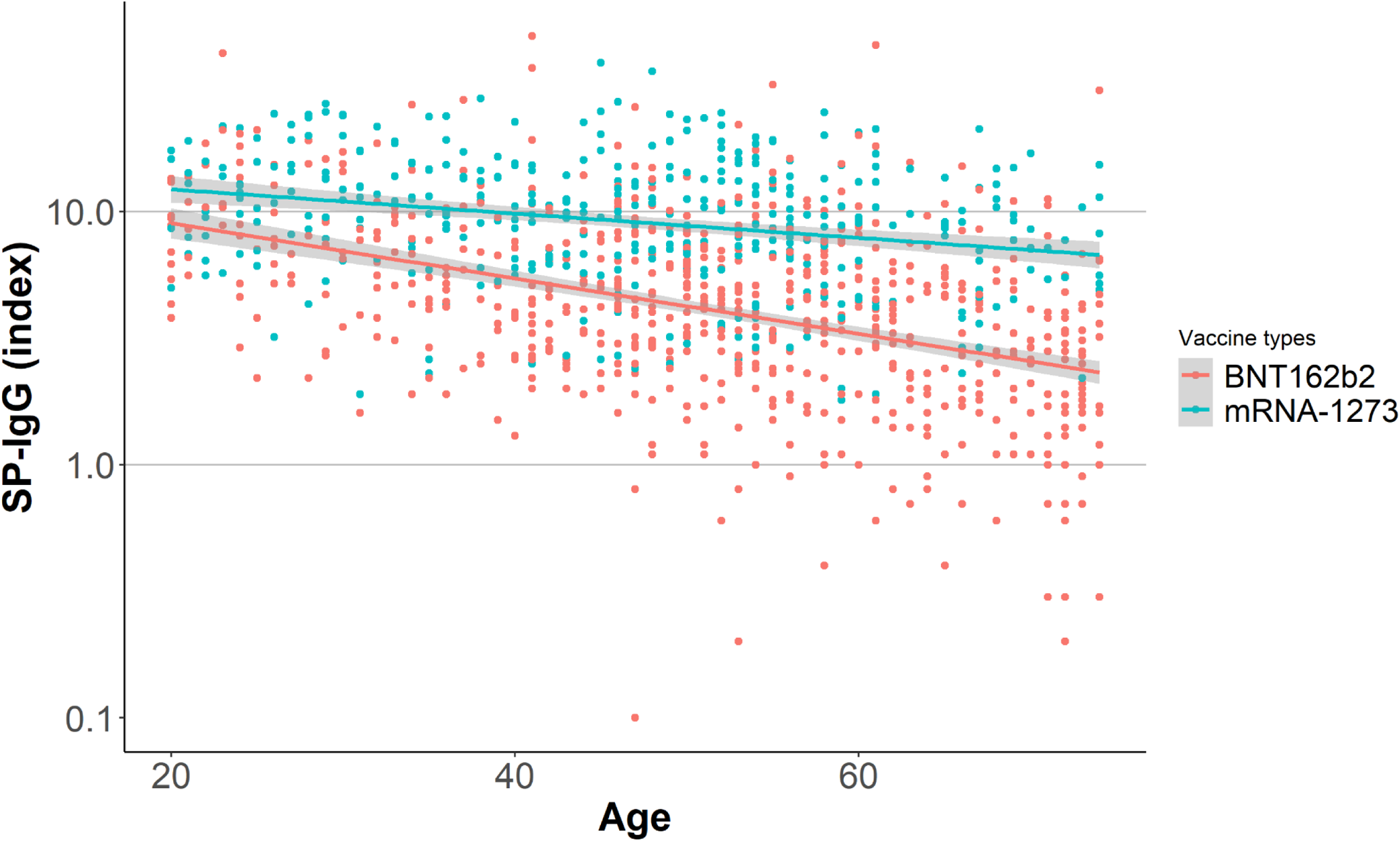
The SP-IgG index according to age and days since the second dose of vaccination by vaccine types (n=1,092) Panel A shows the distribution of SP-IgG index according to age with a regression line and 95% confidence interval band. Panel B shows the distribution of SP-IgG index according to days since the second dose of vaccination with a regression line and 95% confidence interval band.

### SP-IgG index and characteristics among participants with three doses of vaccines

In 76 people with no history of infection and who tested negative for NP-total Ig and received the three doses of vaccines, high SP-IgG index values were observed in those who had received the booster shot at least 7 days before (Figure 3). Approximately 46.1% of these individuals reported that they had the most severe adverse reactions after the third shot (Supplemental Figure 3).

**Figure 3.**
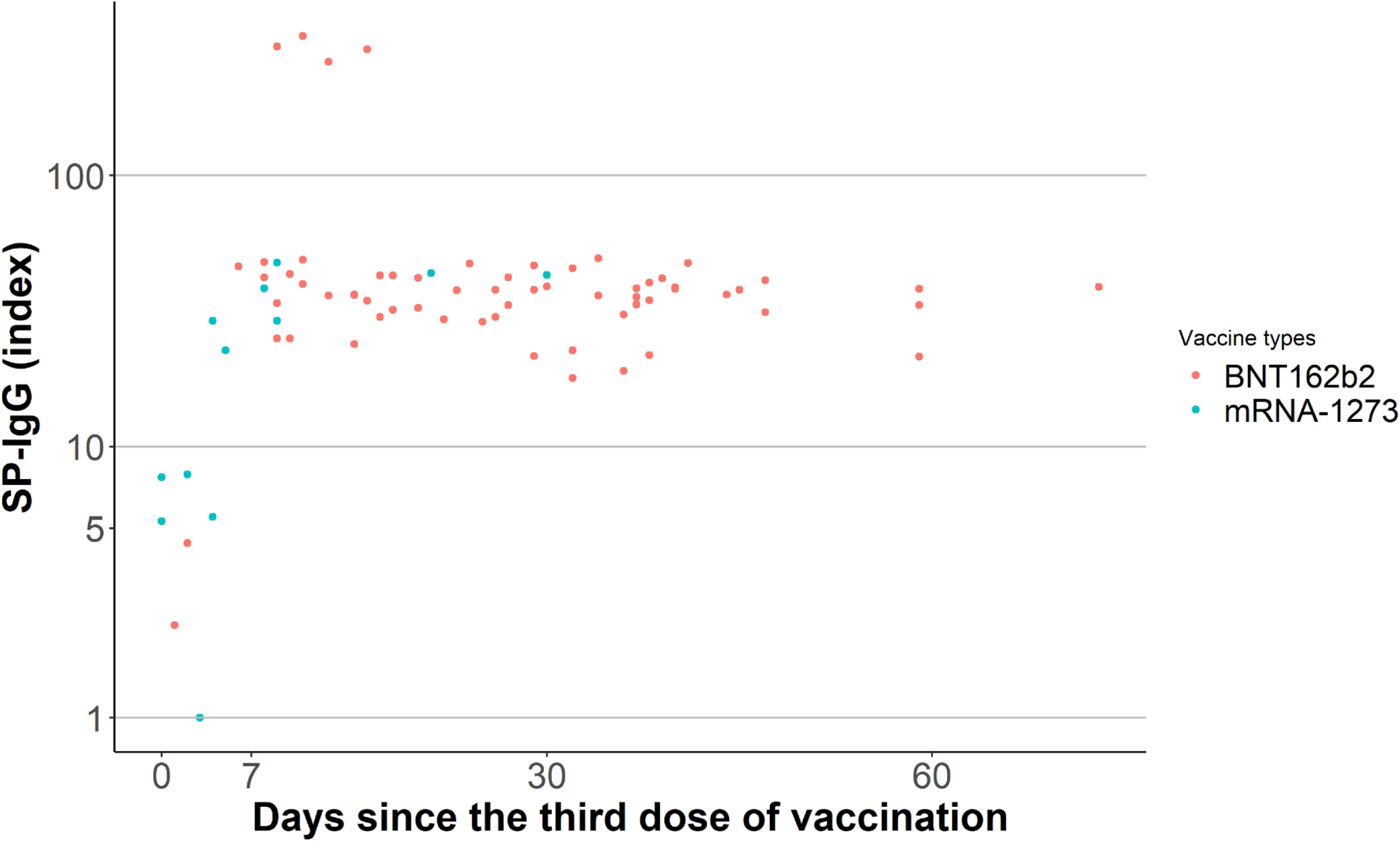
The SP-IgG index according to days since the third dose of vaccination by vaccine types (n=76)

### Prevalence of neutralization antibodies against SARS-CoV-2 variants

Among 123 randomly selected participants, we evaluated the prevalence for neutralization antibodies against the D614G, Delta, Omicron BA.1, and Omicron BA.2 variants. Among these individuals, four were unvaccinated, 115 had received two doses, and four had received three doses of the vaccines. Approximately 87% had neutralizing antibodies to the reference D614G strain, 74% to the Delta strain, 28% to the Omicron BA.1, and 28% to the Omicron BA.2 (Table 2). Among them, the SP-IgG index values were higher among those with positive D614G neutralization antibodies (Supplemental Figure 4) and among those with positive Delta neutralization antibodies (Supplemental Figure 5).

**Table 2.** Prevalence of neutralizing antibodies against D614G, Delta, Omicron BA.1, and Omicron BA.2.

|  | D614G | Delta | Omicron BA.1 | Omicron BA.2 |
| --- | --- | --- | --- | --- |
| **Random subsample, N** | 123 | 123 | 123 | 123 |
| N with positive results | 107 | 91 | 34 | 34 |
| % (95% CI) | 87.0 (79.7–92.4) | 74.0 (65.3–81.5) | 27.6 (20.0–36.4) | 27.6 (20.0–36.4) |
| **Vaccinated two doses and**<br>**SP-IgG index of $\geq 10$ in random subsample, N** | 24 | 24 | 24 | 24 |
| N with positive results | 24 | 24 | 17 | 17 |
| % (95% CI) | 100.0 (85.8–100.0) | 100.0 (85.8–100.0) | 70.8 (48.9–87.4) | 70.8 (48.9–87.4) |
| **Vaccinated three doses, N\*** | 66 | 66 | 66 | 66 |
| N with positive results | 66 | 66 | 66 | 66 |
| % (95% CI) | 100.0 (94.6–100.0) | 100.0 (94.6–100.0) | 100.0 (94.6–100.0) | 100.0 (94.6–100.0) |

Abbreviation: CI, confidence interval.
\*A total of 66 participants who received the booster shot of vaccine against SARS-CoV-2 at least 7 days before were analyzed, after excluding those with prior COVID-19 diagnosis or positive NP-total Ig the analysis.

Furthermore, the SP-IgG index values were higher among those with positive Omicron BA.1 neutralization antibodies (GMT: 14.5; 95% CI: 11.7–18.0) than among those with negative results (GMT: 3.0; 95% CI: 2.4–3.7) (Figure 4A); similar results were observed for BA.2 (GMT in positive Omicron BA.2 neutralization antibodies: 14.5; 95% CI: 11.7–18.0; GMT in negative Omicron BA.2 neutralization antibodies: 3.0; 95% CI: 2.4–3.7) (Figure 4B). In addition, among 66 participants with no history of infection and who tested negative for NP-total Ig and received the third dose of vaccine at least 7 days before, 100% had neutralizing antibodies against the Omicron BA.1 and BA.2 variants (Table 2).

**Figure 4.**
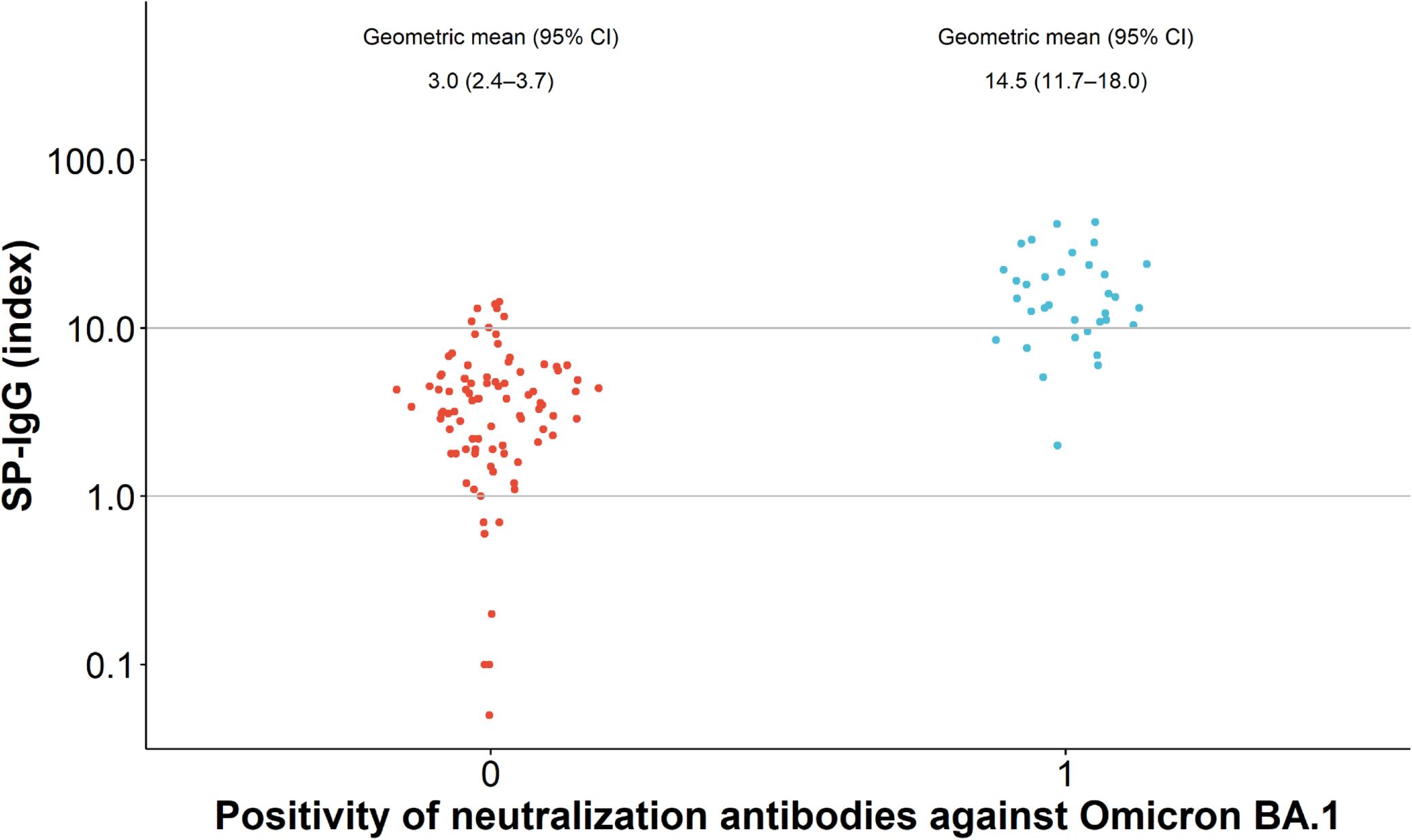
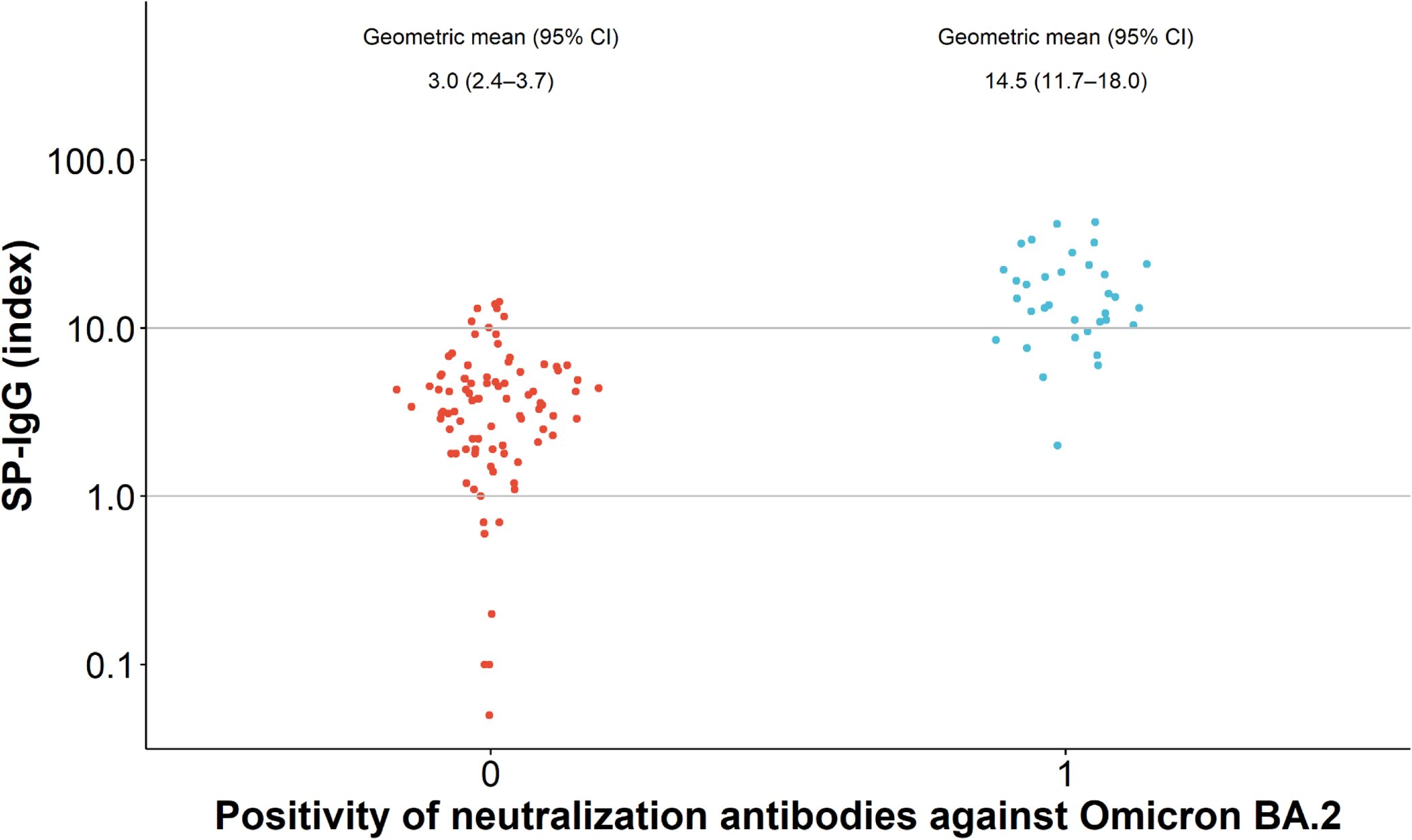
The SP-IgG index according to positivity of neutralization antibodies against Omicron BA.1 and BA.2 variants (n=123) Panel A shows the distribution of SP-IgG index according to positivity of neutralization antibodies against Omicron BA.1 variant. Panel B shows the distribution of SP-IgG index according to positivity of neutralization antibodies against Omicron BA.1 variant. A label of 1 indicates a positive for neutralization antibodies, while a label of 0 indicates a negative for neutralization antibodies. Abbreviation: CI, confidence interval.

## DISCUSSION

In this population-based cross-sectional study with a well-characterized population from Yokohama City as the participants, population-level humoral response to the SARS-CoV-2 and its immune escape variants were assessed at periods when approximately 80% of the total population in Japan have received at two doses of vaccines against SARS-CoV-2. We found that the overall SP antibody positivity rate was 94% during a period from January 30 to February 28, 2022. Moreover, the overall prevalence of the neutralizing antibodies to the Omicron BA.1 and BA.2 variants were 28 % and 28%, respectively. All participants who received the third vaccination at least 7 days before had positive neutralizing antibodies against the Omicron BA.1 and BA.2 variants. Our findings highlight that the population-level immune response against the conventional variant of SARS-CoV-2 is estimated to be high, while the immunity against the Omicron BA.1 and the newly emerged BA.2 variants is only achieved in approximately 28% of the population. The population-level insufficient humoral immunity against the Omicron variants may explain the spread of infection in the sixth wave of the virus during this period in Japan.

Our findings showing that only 28% of our population had neutralization antibodies to both the Omicron BA.1 and BA.2 variants suggest that the second dose of vaccination is not sufficient in most individuals to have neutralization antibodies to these variants. However, our findings also suggest that people who received two doses of vaccines and have high SP-IgG index values tend to have antibodies to the BA.1 and BA.2 variants (Figure 4; Table 2). The same rates of having neutralizing antibodies against the BA.1 and BA.2 variants observed in our study are in line with a recent study showing that the titers of the neutralizing antibodies against the BA.1 and BA.2 variants were similar [20]. Because all participants who received the third vaccination at least 7 days before had positive neutralizing antibodies against the BA.1 and BA.2 variants, the positive rate against BA.1 and BA.2 would be expected to increase if the proportion of those who received three doses of the vaccine increases. However, as vaccine effectiveness against the BA.1 strain is reported to substantially decrease several months after the third vaccination[5], the booster shot of the currently available vaccine alone may not tackle the newly emerged variants of SARS-CoV-2.

Our findings show that 94% of participants had SP-IgG antibodies, which is largely consistent with the fact that approximately 96% of our population had completed their second or third dose of vaccination, 2.8% had been diagnosed with COVID-19, and antibody titers are known to decline over time[21]. Because approximately 87% of the residents aged 20–79 in Yokohama City received the second or third dose of vaccination by the end of February 2022 [22], the proportion of unvaccinated population may be underestimated in our population, suggesting that seroprevalence against emerging variants of SARS-CoV-2 may be even lower than our estimates.

The strengths of this study include its population-based design with a relatively large sample size and detailed assessments of the demographic data, vaccination status, and previous diagnosis of COVID-19. Furthermore, we measured SP-IgG and NP-total Ig using highly accurate quantitative measurements. Additionally, we assessed the population seroprevalence of the neutralizing antibodies against the variants including Omicron BA.2. However, the present study has several limitations. First, the cross-sectional nature of the study limits the ability to investigate causality between measured variables. However, given the growing evidence regarding the effectiveness of vaccination on the immunity against and risk of contracting COVID-19, our findings provide an accurate description of the humoral immunity against variants, including assessments of the emerging variants such as Omicron BA.1 and BA.2. Second, the assay used to assess the positivity of the neutralization antibodies was qualitative. That is, we could only evaluate the presence or absence of the neutralization antibodies at a certain threshold, equivalent to the neutralizing titer of 51 or more in the HIV-based pseudovirus method [15]. However, our findings are consisitent with published studies using the quantitative assessments of the neutralization antibodies[23, 24]. Third, the response rate of our study was approximately 21%, which is not sufficiently high. However, to adjust the differences in age and sex distribution between our study and the overall population of Yokohama City, weighted prevalence rates were estimated and the results were almost identical. Lastly, our study provides evidence regarding a certain period of time in Yokohama City, Japan, and the generalizability may be limited. However, because the vaccination has been similarly provided to the nationwide population in Japan and the vaccination rates in Japan are similar to other high-income countries[10], the findings could be generalized to other populations.

To summarize, in this population-based cross-sectional study conducted in Japan, the majority of participants had SP-IgG antibodies, and the older they were and the more the number of days that had elapsed after the second vaccination, the lower their antibody titers tended to be. The overall prevalence of the neutralizing antibodies to Omicron BA.1 and BA.2 variants were only 28% from January to February 2022 in Japan; however, when more than 7 days had elapsed since the third dose of vaccination, the neutralizing antibody possession rates against the Omicron BA.1 and BA.2 variants were 100%. Due to the threat of the BA.2 variant becoming dominant globally, these findings have certain implications.

## Data Availability

All data produced in the present study are available upon reasonable request to the authors.

## ACKNOWLEDGEMENTS

We would like to acknowledge all the participants for their participation in this research. We thank Sho Katsuragawa, Toshiya Sakota, and the staff members at the Yokohama City University for their assistance. We also thank Mr. Mitsuru Kurata at the Prime Health Partners Co., Ltd. for his contribution to this study.

## Funding

This work was supported by the grant for 2021–2022 Strategic Research Promotion (No.SK202116) of Yokohama City University. The work was also supported by the Japan Agency for Medical Research and Development (AMED) under Grant Number: JP21fk0108104.

## Conflict of Interest Statement

SY is an employee of Integrity Healthcare Co., Ltd. NO is an employee of Tosoh Corporation. These companies did not have any role in study design, data collection and analysis, decision to publish, or preparation of the manuscript. HK received grants from Shionogi & Company, Limited, and Asahi Kasei Pharma & Co., Inc. Other authors stated no conflict of interest.

## Figure legends

**Supplemental Figure 1.**
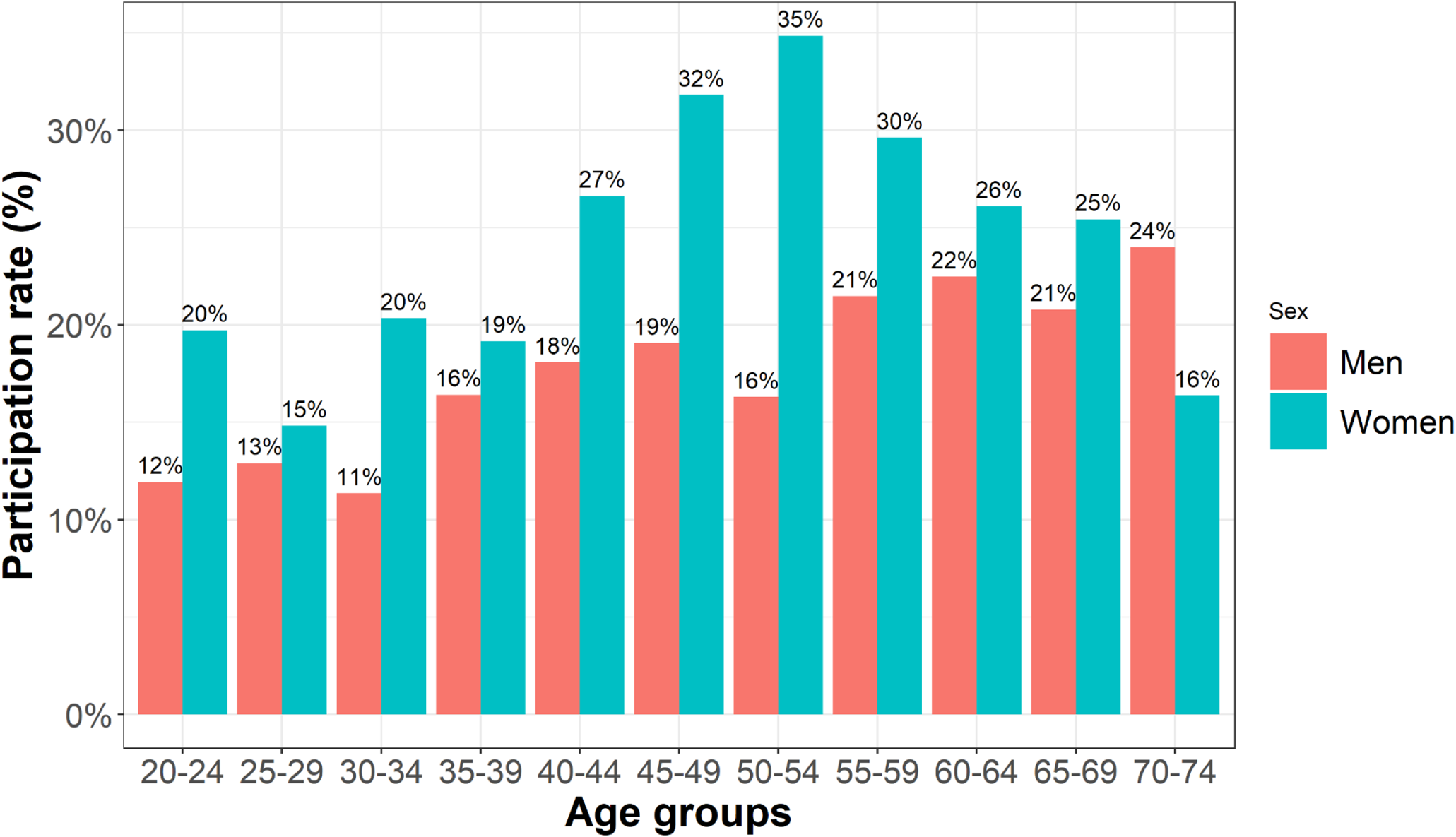
Participation rate according to age groups.

**Supplemental Figure 2.**
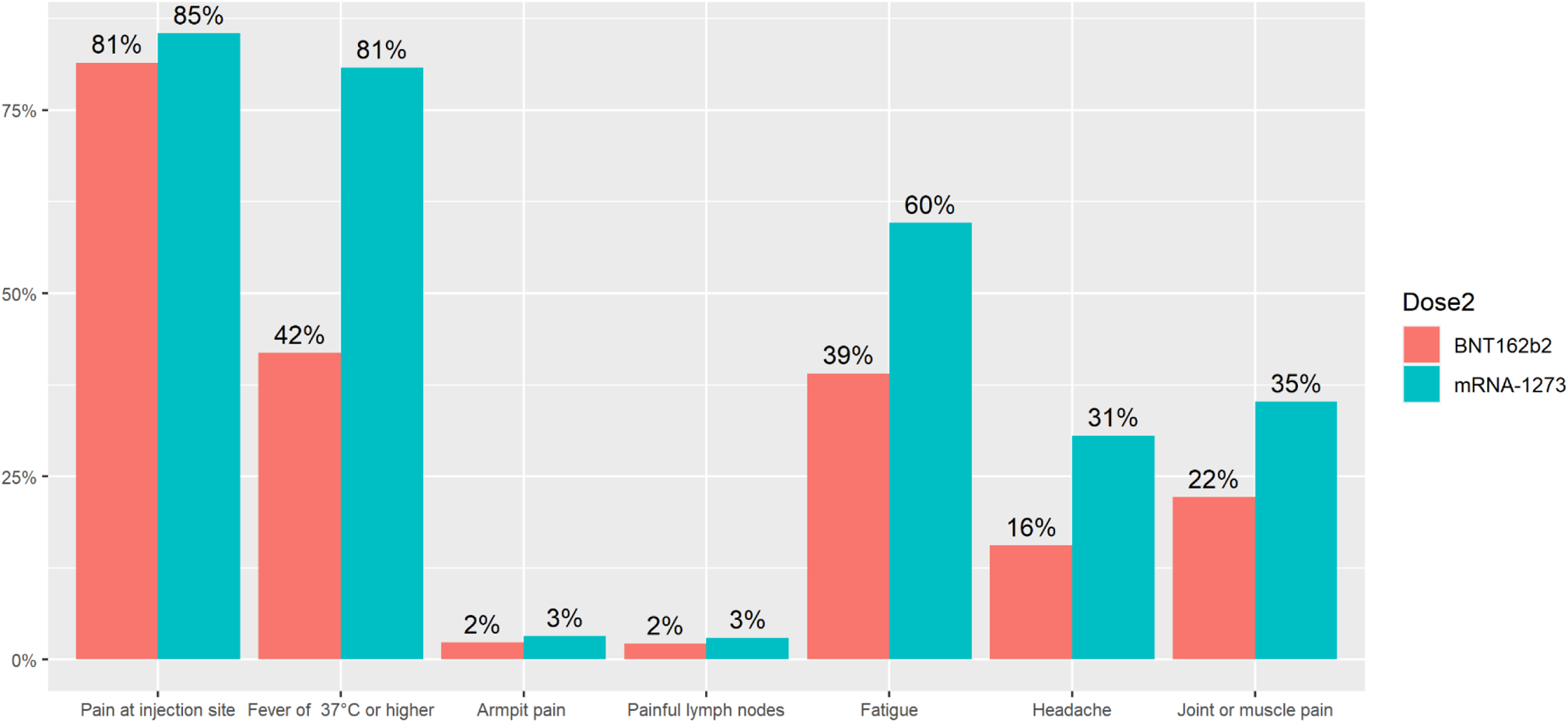
Adverse reactions after the two doses of vaccines

**Supplemental Figure 3.**
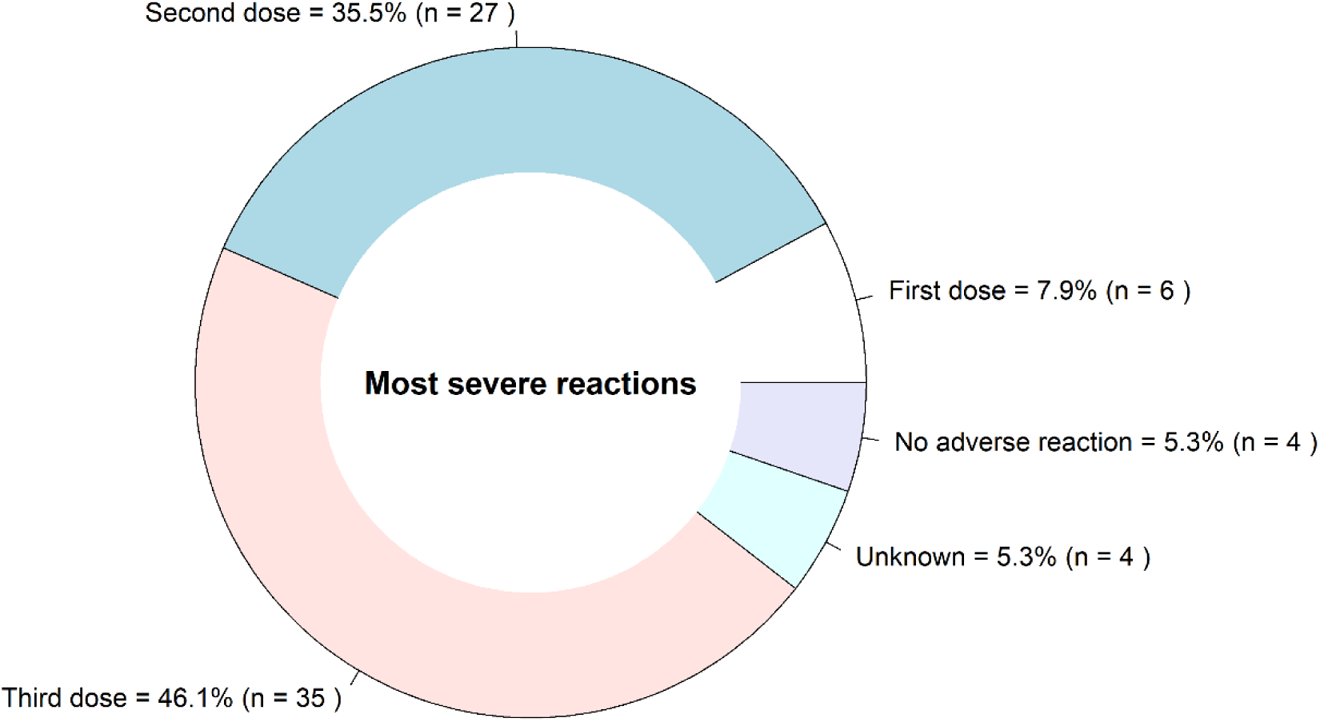
The dose at which most severe adverse reactions were experienced among those received three doses of vaccines.

**Supplemental Figure 4.**
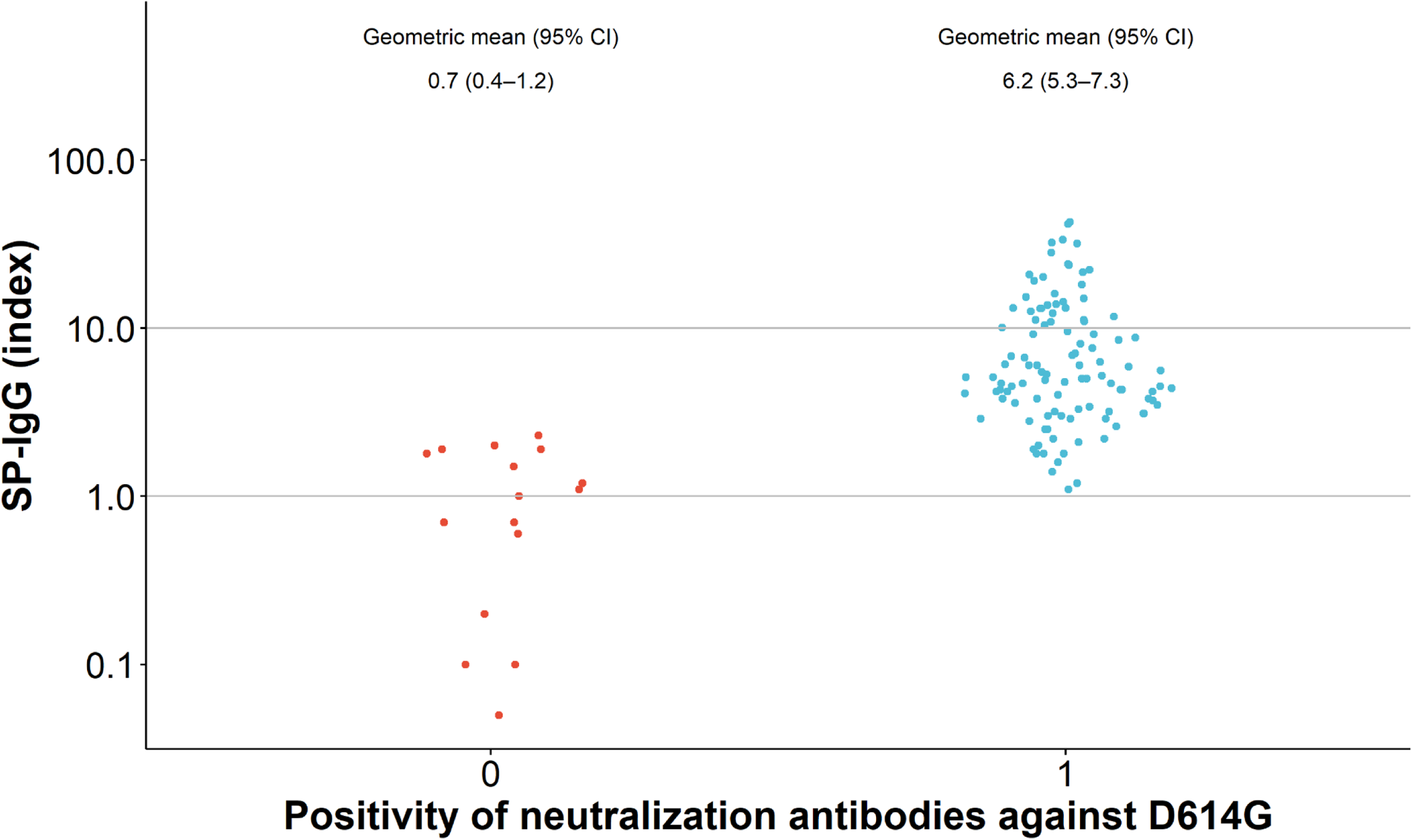
SP-IgG index according to positivity of neutralization antibodies against D614G.

**Supplemental Figure 5.**
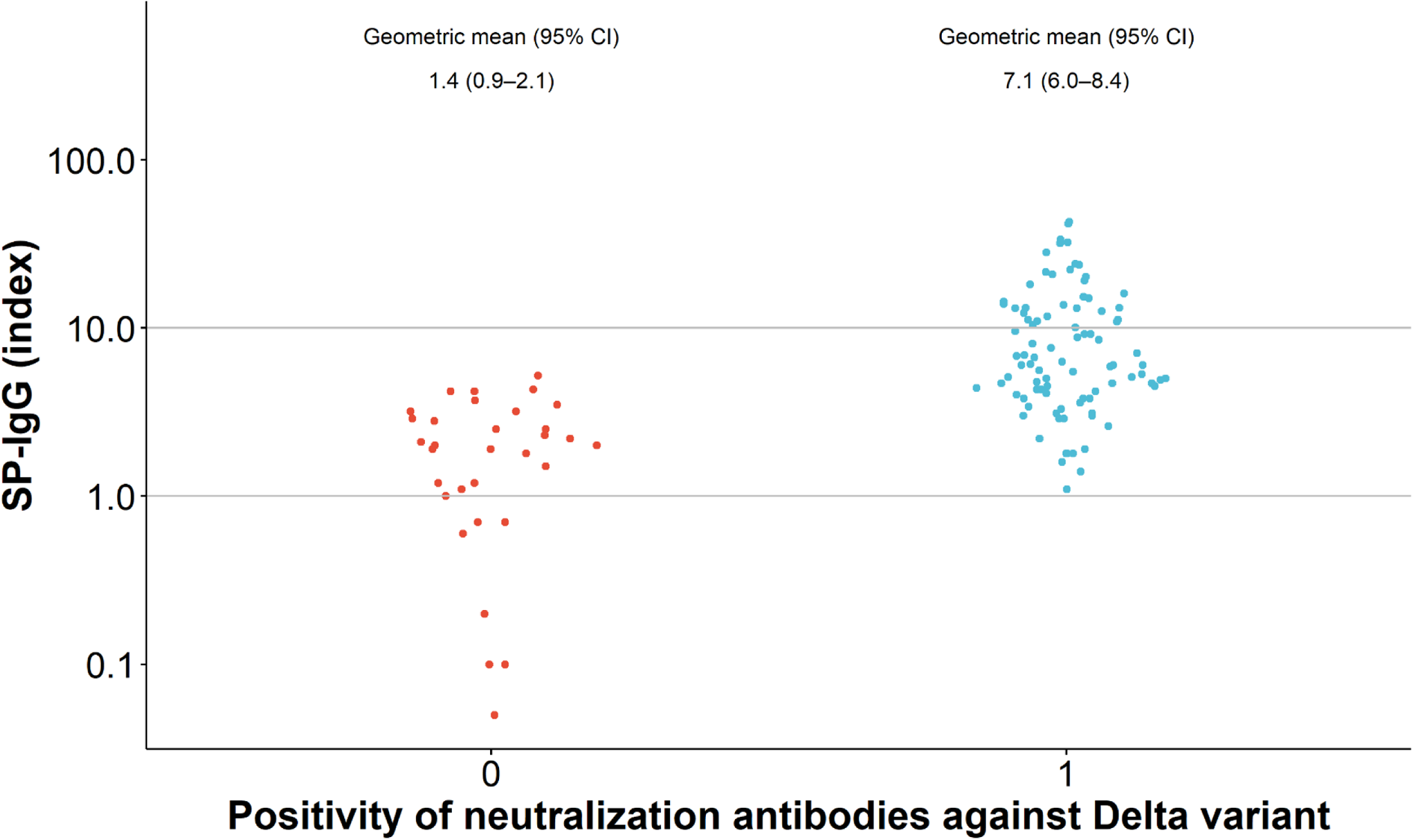
SP-IgG index according to positivity of neutralization antibodies against Delta variant.

## Notes

Conflict of Interest SY is an employee of Integrity Healthcare Co., Ltd. NO is an employee of Tosoh Corporation. These companies did not have any role in study design, data collection and analysis, decision to publish, or preparation of the manuscript. HK received grants from Shionogi & Company, Limited, and Asahi Kasei Pharma & Co., Inc. Other authors stated no conflict of interest.

Funding Statement This work was supported by the grant for 2021–2022 Strategic Research Promotion (No.SK202116) of Yokohama City University. The work was also supported by the Japan Agency for Medical Research and Development (AMED) under Grant Number: JP21fk0108104.

### Author Declarations

The IRB of Yokohama City University gave ethical approval for this work.

## REFERENCES

1. The Lancet. Access to COVID-19 vaccines: looking beyond COVAX. Lancet 2021; 397:941.

2. Yamamoto S, Maeda K, Matsuda K, et al. COVID-19 breakthrough infection and post-vaccination neutralizing antibody among healthcare workers in a referral hospital in Tokyo: a case-control matching study. Clin Infect Dis 2021; ciab1048.

3. Bergwerk M, Gonen T, Lustig Y, et al. Covid-19 breakthrough infections in vaccinated health care workers. N Engl J Med 2021; 385:1474–84.

4. Prime Minister’s Office of Japan. COVID-19 vaccines. Available at: https://japan.kantei.go.jp/ongoingtopics/vaccine.html. Accessed March 19th 2022.

5. Andrews N, Stowe J, Kirsebom F, et al. Covid-19 Vaccine Effectiveness against the Omicron (B.1.1.529) Variant. N Engl J Med 2022; DOI: 10.1056/NEJMoa2119451.

6. Planas D, Saunders N, Maes P, et al. Considerable escape of SARS-CoV-2 Omicron to antibody neutralization. Nature 2022; 602:671–5.

7. Lu L, Mok BW, Chen LL, et al. Neutralization of SARS-CoV-2 Omicron variant by sera from BNT162b2 or Coronavac vaccine recipients. Clin Infect Dis 2021; DOI: 10.1093/cid/ciab1041.

8. The Johns Hopkins Coronavirus Resource Center. Daily confirmed new cases (7-day moving average). Available at: https://coronavirus.jhu.edu/data/new-cases. Accessed 3/26 2022.

9. The Our World in Data. Share of SARS-CoV-2 sequences that are the omicron variant. Available at: https://ourworldindata.org/covid-cases. Accessed 03/26 2022.

10. The Johns Hopkins Coronavirus Resource Center. Vaccination progress across the world. Available at: https://coronavirus.jhu.edu/vaccines/international. Accessed 3/26 2022.

11. Madhi SA, Kwatra G, Myers JE, et al. Population Immunity and Covid-19 Severity with Omicron Variant in South Africa. New England Journal of Medicine 2022.

12. Ren Z, Nishimura M, Tjan LH, et al. Large-scale cross-sectional seroepidemiologic study of COVID-19 in Japan: Acquisition of herd immunity and the vaccines’ efficacy. medRxiv 2022; DOI: 10.1101/2022.01.13.22269203.

13. World Health Organization. Weekly epidemiological update on COVID-19 - 22 March 2022. Available at: https://www.who.int/docs/default-source/coronaviruse/situation-reports/20220322_weekly_epi_update_84.pdf?sfvrsn=9ec904fc_4&download=true. Accessed 03/25 2022.

14. Kato H, Miyakawa K, Ohtake N, et al. Antibody titers against the Alpha, Beta, Gamma, and Delta variants of SARS-CoV-2 induced by BNT162b2 vaccination measured using automated chemiluminescent enzyme immunoassay. J Infect Chemother 2022; 28:273–8.

15. Miyakawa K, Stanleyraj JS, Kato H, et al. Rapid detection of neutralizing antibodies to SARS-CoV-2 variants in post-vaccination sera. J Mol Cell Biol 2021; 13:918–20.

16. Goto A, Go H, Miyakawa K, et al. Sustained neutralizing antibodies 6 months following infection in 376 Japanese COVID-19 survivors. Front Microbiol 2021; 12:661187.

17. Kubo S, Ohtake N, Miyakawa K, et al. Development of an automated chemiluminescence assay system for quantitative measurement of multiple anti-SARS-CoV-2 antibodies. Front Microbiol 2020; 11:628281.

18. Miyakawa K, Kubo S, Stanleyraj Jeremiah S, et al. Persistence of Robust Humoral Immune Response in Coronavirus Disease 2019 Convalescent Individuals Over 12 Months After Infection. Open forum infectious diseases 2021; 9:ofab626-ofab.

19. Lumley T. Analysis of Complex Survey Samples. Journal of Statistical Software 2004; 9:1–19.

20. Yu J, Collier A-rY, Rowe M, et al. Neutralization of the SARS-CoV-2 Omicron BA.1 and BA.2 Variants. New England Journal of Medicine 2022.

21. Kato H, Miyakawa K, Ohtake N, et al. Vaccine-induced humoral and cellular immunity against SARS-CoV-2 at 6 months post BNT162b2 vaccination. medRxiv 2021; DOI: 2021.10.30.21265693.

22. The City of Yokohama. Number of vaccination according to age groups and dates. Available at: https://www.city.yokohama.lg.jp/kurashi/kenko-iryo/yobosesshu/vaccine/wakuchinsessyudeta.files/141003_yokohama_covid19_numerator.csv. Accessed March 22th 2022.

23. Iketani S, Liu L, Guo Y, et al. Antibody evasion properties of SARS-CoV-2 Omicron sublineages. Nature 2022.

24. Kawaoka Y, Uraki R, Kiso M, et al. Characterization and antiviral susceptibility of SARS-CoV-2 Omicron/BA.2. Res Sq 2022.

